# Inferior vena cava ultrasound *versus* passive leg raising test in guiding fluid administration in surgical patients prior to spinal anaesthesia: a post-hoc analysis of the ProCRHYSA randomized trial

**DOI:** 10.1101/2023.05.22.23290367

**Authors:** Samuele Ceruti, Andrea Glotta, Mathieu Favre, Edoardo Tasciotti, Giovanni Bona, Antonietta Petrusic, Alain Borgeat, José Aguirre, Andrea Saporito

## Abstract

**Background:** Spinal anaesthesia is commonly used for many surgical procedures. One of its potential complications is arterial hypotension, which is nowadays prevented by an empirical fluid administration without any hemodynamic status assessment. However, this practice could increase the risk of volume overload in cardiovascular high-risk patients. Two non-invasive tests are performed to identify fluid-responsiveness: the Inferior Vena Cava Ultrasound (IVCUS) and the Passive Leg Raising Test (PLRT). Aim of this post-hoc analysis was to compare these two methods in spontaneous-breathing patients to assess fluid responsiveness before spinal anaesthesia. Primary outcome was to analyze the incidence of arterial hypotension after spinal anaesthesia in elective surgery patients. Secondary endpoints compared the total fluids amount, the vasoactive drugs administered and the time needed to accomplish the whole procedure in both groups.

**Results:** The patients analyzed were 132 in the IVCUS group and 148 in the PLRT group; 39.6% of all patients developed arterial hypotension after spinal anaesthesia, 34.8% in the IVCUS group and 43.9% in the PLRT group (Chi-square 2.39, df = 1, p = 0.77). The mean total fluids amount was 794 ± 592 ml; 925 ± 631 ml for IVCUS group and 678 ± 529 ml for PLRT group (p < 0.001). Patients needed vasoactive drugs to restore normal arterial pressure were 18.2% of total, 15% in the IVCUS group and 20% in the PLRT group (p = 0.136). The mean time required to complete the entire procedure was 52 ± 18 min, 48 ± 10 min in the IVCUS group and 56 ± 13 min in the PLRT group (p < 0.001). Complications or out of protocol treatment were registered in 4.6% patients.

**Conclusions:** Fluid responsiveness assessment in spontaneous breathing patients before spinal anaesthesia could potentially prevent the risk of post-spinal hypotension. In elective surgery, IVCUS could be an accurate method to guide fluid administration in patients undergoing spinal anaesthesia, reducing the incidence of post-spinal hypotension when compared to PLRT.

## INTRODUCTION

Spinal anaesthesia is commonly used for a wide range of surgical procedures [1]. Despite its routine use, it remains a technique burdened by potentially severe complications, especially arterial hypotension [2–8]. Subarachnoid administration of local anaesthetics blocks the autonomous nervous system controlling vascular tone [4,6], potentially leading to a sudden decrease in peripheral vascular resistances which may result in arterial hypotension, further aggravated by a relative hypovolemia [2,7–11]. Transient hypotensive episodes are generally well tolerated in patients without severe comorbidities, due to cardiovascular compensatory mechanism [11,12]. However, these events may lead to major complications in emergency situations and in cardiovascular high-risk patients [5,10]; patients older than 65 years [2,13] as well as with an ongoing drugs therapy such as beta-blockers [2], angiotensin-converting enzyme (ACE)-inhibitors, selective serotonin reuptake inhibitors (SSRIs) or monoamine oxidase (MAO) inhibitors [14] presented an increased risk of developing a severe hypotensive response after spinal anaesthesia. The induced reduction in both cardiac output and systemic vascular resistance (SVR) are the main causes of the hypotension. Furthermore, Monk et al. observed how a significant intra-operatory hypotension is associated with a greater mortality in the 30 days following surgery [15]. Spinal anaesthesia related mortality may thus be currently still underestimated.

In common clinical practice, post-spinal hypotension is usually prevented by an empirical fluid administration [3–5,10,16], without a routine evaluation of patient’s hemodynamic status. However, this practice brings an intrinsic risk of volume overload, which may be counterproductive in some high-risk subjects, as mentioned above [5,16,17].

In this setting, two main non-invasive tests have been developed to identify fluid-responsiveness in spontaneously breathing patients: the Inferior Vena Cava Ultrasound (IVCUS) and Passive Leg Raising Test (PLRT) [18,19]. Nowadays ultrasound evaluation, especially cardiac ultrasonography, has become widely used in the clinical practice and in 2013 the American Society of Echocardiography (ASE) published a consensus suggesting the use of *Point of Care UltraSound* (POCUS) as adjunctive therapeutic aid at patients’ bedside [20]. Following these recommendations, the use of POCUS is progressively increasing in the preoperative setting to identify and prevent complication before spinal or general anaesthesia [21–23]. The IVCUS consists in a measurement of the IVC diameter variation by M-mode ultrasound analysis during spontaneous breathing activity [24,25]; regarding this technique, while recent data in the non-critical patients population are encouraging [26,27], there are contrasting results in critically ill patients, for whom the method had originally being validated [18,25,26,28–32].

The PLRT consists in passively raising the patient’s legs to increase venous return and therefore cardiac output [19]; potential fluid responsiveness can consequently be assessed by measuring changes in stroke volume (SV) with echocardiography or other non-invasive methods, like etCO_2_ [16,19,33,34]. PLRT has been identified as a test for predicting fluid responsiveness in spontaneous breathing patients [35], and it has been recommended by the European Society of Intensive Care Medicine (ESICM) as a potential non-invasive method to analyze fluid-responsiveness in critically ill patients [36].

We have previously published the protocol of the *ProCRHYSA trial* [37], aiming at determining whether these two methods (IVCUS and PLRT) are effective in guiding fluid therapy to reduce both the hypotension rate and the amount of volume needed in non-critical patients undergoing elective spinal anaesthesia for surgical procedures. We previously concluded that IVCUS resulted a valid method to significantly reduce the post-anaesthesia hypotension rate in comparison to the standard of care, through the identification of those patients who are eligible for fluid replacement [37]. Some studies have identified the PLRT as the preferred method for fluid-responsive identification in spontaneous-breathing patients [27,35,38]; however, the aforementioned ProCRHYSA trial indirectly suggested a relatively little benefit of PLRT use in comparison to standard of care [39]. The main objective of this study is the comparison between these two non-invasive techniques to determine fluid-responsiveness in the context of preoperative spinal anaesthesia.

## MATERIAL AND METHODS

### 1. Study design and enrollment criteria

The analyses of this project are based on the data collected in the ProCRHYSA trial [37], which was a prospective, controlled, randomized monocentric clinical trial including consecutive pre-operative patients receiving elective spinal anaesthesia, treated according to current clinical practice or with optimized fluid-responsiveness status assessment according to IVCUS o PLRT methods [37]. The trial was performed in a tertiary care hospital (Bellinzona Regional Hospital - ORBV, Service of Anaesthesia, Bellinzona) in Switzerland.

Inclusion criteria were adult patients of both sexes, with an anaesthesia risk score I to III on the basis of the American Society of Anaesthesiology (ASA) scoring system [40], undergoing an elective surgical intervention under spinal anaesthesia. Exclusion criteria were the need of invasive blood pressure monitoring, any condition determining a pre-procedural arterial hypotension, patients with unilateral anaesthetic block, patients needing any form of assisted ventilation prior or during the surgical intervention and those unable to give informed consent [37] (Table 1).

**TABLE 1:** inclusion and exclusion criteria

| Inclusion | Exclusion |
| --- | --- |
| Adults from 18 to 80 years | Patients requiring invasive blood pressure monitoring (arterial catheter, pulmonary catheter, thermodilution catheter) |
| Both sexes | Patients with pre-procedural hypotension |
| Not critical patients undergoing spinal anaesthesia for an elective surgery<br>ASA class from I to III | Patients with unilateral anaesthetic block<br>Patients needing assisted ventilation before or during the surgical intervention |
|  | Patients unable to give informed consent |
|  | Contraindications to perform spinal anaesthesia (previous back surgery in the lumbar region, clinical high-risk conditions like thrombocytopenia < 50 G/L or coagulation abnormalities) |
|  | Obstetrical population |

Eligibility inclusion/exclusion criteria from the ProCRHYSA trial.

The CONSORT reporting guidelines has been used throughout the entire trial [41] (SM 1); all clinical and pharmacological data were registered and collected in a codified electronic database by an anaesthesiologist not directly involved in patient management.

### 2. Clinical trial conduction

The study started upon patient’s arrival in the operating room (time 0) and ended 30 minutes after completion of spinal anaesthesia. It was divided in the *pre-anaesthetic phase* from time 0 to the beginning of spinal anaesthesia, in the *anaesthetic phase* corresponding to the performance of spinal anaesthesia, and in the *post-anaesthetic phase* from the end of anaesthesia to the following 30 minutes. Spinal anaesthesia was performed following a standardized procedure, with intrathecal administration of hyperbaric bupivacaine 0.5%, with dosing according to surgical procedure and patient’s body structure, at L_3_-L_4_ level in the lateral decubitus position, using a 27G pencil point cranially orientated spinal needle (BBraun Medical SA, Melsungen, Germany). After the injection patients rested supine during 30 minutes before surgery. A nurse not involved in the trial and blind to groups allocation was assigned to assess the sensory block extension with cold test for a T_H8_-T_H6_ targeted level block [39]. Arterial hypotension was previously defined as two measurements of systolic arterial pressure inferior to 90 mmHg, a mean arterial pressure (MAP) inferior to 60 mmHg [27,39], any fall in systolic blood pressure more than 50 mmHg or more than 25% from baseline, a reduction in mean arterial pressure of more than 30% from baseline and/or clinical signs and symptoms of inadequate perfusion [27,37]. After spinal anaesthesia, hypotensive patients were treated according to a standard protocol with crystalloid replacement and vasoactive drugs (e.g. ephedrine, phenylephrine or atropine depending on the clinical status and in agreement with the anaesthesiologist’s evaluation), until haemodynamic stability was accomplished and all clinical criteria for arterial hypotension were solved.

#### 2.1 IVCUS group

Patients allocated in this group underwent IVCUS-based fluid status assessment before spinal anaesthesia. According to the protocol, an *IVC collapsibility index* (IVC-CI) greater than 36% was chosen as cut-off [24–26,37,39,42]; the index was measured in the IVC intra-abdominal portion (2 cm from the right atrium) through a subcostal view and was calculated as (IVC_max_-IVC_min_)/IVC_max_, with the diameters being registered in spontaneous breathing activity for 30 seconds. An infusion of 500 ml of crystalloid for 20 minutes was considered adequate and safe, although no published consensus regarding this matter exists [16]. Patient’s haemodynamic status was re-assessed after each refilling: patients with an IVC-CI greater than 36% were identified as *responsive patients*: crystalloid fluids were therefore administered, followed by IVCUS reassessment; otherwise, they were considered as *unresponsive patients* and directly underwent spinal anaesthesia.

#### 2.2 PLRT group

Patients in this group underwent PLRT prior to spinal anaesthesia. Active leg elevation exerts an orthosympathetic reflex that can increase cardiac output; passive lower limb test had the advantage of mobilizing lower limb venous blood (about 300-500 ml) without activating the orthosympathetic reflex, making it possible to quantify the clinical response after a bolus of fluids [37]. Before the test, patients waited 5 minutes in a semi-recumbent supine position at 45°, to reach a phase of hemodynamic and respiratory balance and general stability, as well as to reduce anxiety and stress. The etCO_2_ measurement with continuous CO_2_ sampling through nasal cannula was performed before and after PLRT, while patients kept a semi-recumbent position at 45°. Patients with an etCO_2_ increase of more than 12% from baseline were classified as *responsive* [11], otherwise they were classified as *unresponsive* to fluid administration. Responsive patients received fluids according to the same protocol described for the IVCUS group and were reassessed after each fluid administration through the same PLRT. Spinal anaesthesia was performed once *unresponsiveness* to fluid administration was reached. Follow-up was identical compared with the previous group.

### 3. Outcomes

In this post-hoc analysis of ProCRHYSA trial, the primary endpoint was to report and compare the post-spinal anaesthesia hypotension rate both in the IVCUS group and in the PLRT group. Secondary endpoints were a comparison about total fluids amount (further stratified according to pre/post-anaesthesia stage), about the amount of vasoactive drug use in both groups and about the time needed to accomplish the whole procedure, since the beginning of the pre-anaesthesia phase until the end of the post-anaesthesia phase (further stratified according to pre/post-anaesthesia stage).

### 4. Statistical analysis

Sample size calculation and randomization were described in the original trial [37]. Descriptive statistic was performed to summarize the clinical collected data. According to data distribution, verified by Kolmogorov-Smirnov test and Shapiro–Wilk test, were presented as mean ± SD or median (IQR, min-max) for continuous variables and as absolute number (percentage) for categorical variables. Differences between patients’ outcomes were studied by *t-test* for independent groups or by *Mann-Whitney test* if non-parametric analysis was required. Similarly, comparison of clinical evolution over time was performed by t*-paired test* or by non-parametric *Wilcoxon test*, depending on data distribution. Study of differences between groups of categorical data was carried out by *Chi-square* statistics. All Confidence Intervals (CI) were established at 95%; significance level was established to be < 0.05. Statistical data analysis was performed using the SPSS 26.0 package (SPSS Inc, USA).

### 5. Ethics considerations

The study has been approved by the Ethics Committees (Ref. Cantonal Ethics Committee n. CE2796, international registration number NCT02070276); written informed consent was obtained from patients prior to randomization. There was no funding source for this study.

## RESULTS

### Baseline characteristics

During the study period of the ProCRHYSA trial (3 arms, randomized, parallel group trial), from May 2014 to February 2019, 447 consecutive patients were enrolled and randomized into the tree arms. In this post-hoc analysis, patients enrolled and randomized into the IVCUS and PLRT arms were directly compared (Figure 1); a total of 298 patients (149 for each arm) were recruited, 17 and 1 patients respectively in the IVCUS and PLRT group were subsequently excluded for troubles related to the correct acquisition of the required measures (namely poor ultrasonographic window and intolerance to nasal cannula for etCO_2_ measurement). Therefore, data from 132 patients in the IVCUS group and 148 patients in PLRT group were analyzed (Figure 1). The mean age resulted 56 ± 12 years; 173 (61.7%) patients were men, 244 (87.1%) were classified as ASA I-II and 36 (12.8%) as ASA III. Seventy-five patients (26.8%) resulted on an antihypertensive treatment (14.3% on beta-blockers, 10.7% on ACE-inhibitors, 11.1% on other drugs) and 6.4% were on psychotropic drugs (4.3% on antidepressants, 2.1% on SSRI, while no patients were on MAO-I); pre-anaesthesia hemodynamic data were also compared for blood pressure (p = 0.121), heart rate (p = 0.533) and SpO_2_ (p = 0.04). Clinical data and demographic characteristics of both groups are reported in Table 2.

**FIGURE 1:**
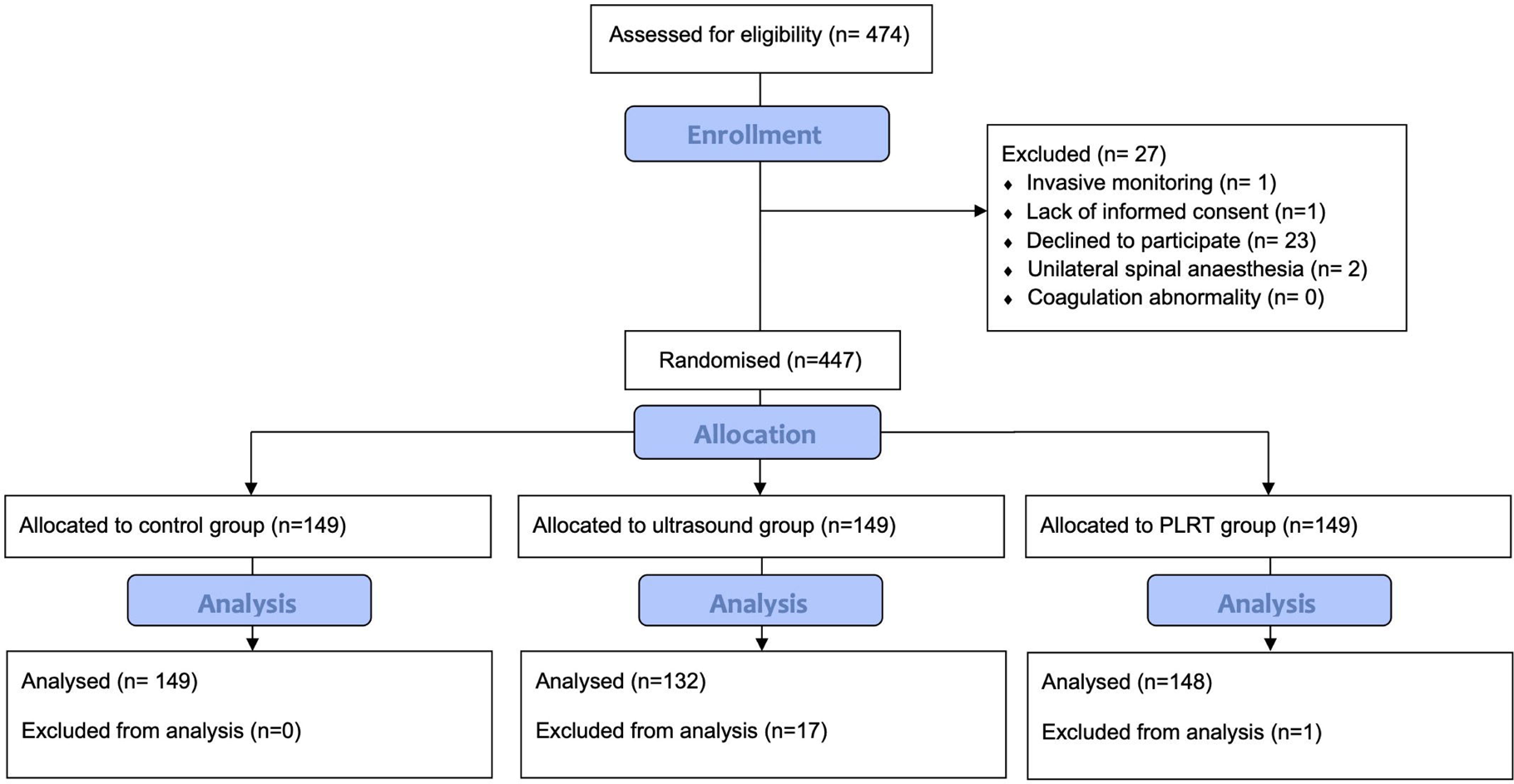
Study flow chart. Flow chart according to CONSORT of the primary study from which the post-hoc analysis was carried out, involving the ultrasound (arm B) and PLRT (arm C) groups.

**TABLE 2:** baseline characteristics

|  | **IVCUS group**<br>**(n = 132)** | **PLRT group**<br>**(n = 148)** | ***p value*** |
| --- | --- | --- | --- |
| Male sex | 82 (62.2%) | 91 (61.5%) |  |
| Age [yrs] | 55.9 ± 18.3 | 57.2 ± 18.3 | 0.192 |
| 18 – 65 | 80 (60.6%) | 88 (59.5%) |  |
| > 65 | 52 (39.4%) | 60 (40.5%) |  |
| Weight [Kg] | 76.7 ± 16.7 | 77.8 ± 15.1 | 0.812 |
| ASA | 1 (1, 1-3) | 1 (1, 1-3) | 0.150 |
| ASA I-II | 114 (86.4 %) | 130 (87.8 %) |  |
| ASA III | 18 (13.3 %) | 18 (12.2 %) |  |
| **Anti-hypertensive therapy** | 37 (28%) | 38 (25.7%) | 0.178 |
| *B-blockers* | 19 (14.4%) | 21 (14.2%) | 0.002 * |
| *ACE-inhibitors* | 16 (12.1%) | 14 (9.5%) | 0.854 |
| *Other* | 16 (12.1%) | 15 (10.1%) | 0.585 |
| **Psychotropic drugs** | 7 (5.3%) | 11 (7.4%) | 0.260 |
| *SSRI* | 4 (3%) | 2 (1.4%) | 0.587 |
| *Anti-depressive* | 3 (2.3%) | 9 (6.1%) | 0.558 |
| *IMAO* | 0 (0%) | 0 (0%) | - |
| **At anesthesia** |  |  |  |
| *Systolic Arterial Pressure*<br>*[mmHg]* | 133.6 ± 21.6 | 137.9 ± 24.6 | 0.500 |
| *Diastolic Arterial Pressure*<br>*[mmHg]* | 71.9 ± 10.9 | 73.5 ± 12.7 | 0.621 |
| *Mean Arterial Pressure*<br>*[mmHg]* | 92.3 ± 13.2 | 95 ± 15.1 | - |
| *Heart Rate [bpm]* | 71.2 ± 12.5 | 70.3 ± 11.5 | 0.670 |
| *SpO<sub>2</sub> [%]* | 96.9 ± 2.2 | 97.4 ± 1.8 | 0.956 |

Baselines patients' clinical characteristics according to randomization into IVCUS and PLRT group.
All data are reported as mean (SD) or median (IQR, min – max) according to Kolmogorov-Smirnov
test data distribution.

### Primary endpoint

After spinal anaesthesia, 111 (39.6%) of all patients developed arterial hypotension; the hypotension rate resulted 34.8% (46 patients) in the IVCUS group and 43.9% (65 patients) in the PLRT group (Chi-square 2.39, df = 1, p = 0.77); the odds ratio (OR) for patients pre-treated according to IVCUS test compared to PLRT was 0.68 (95% CI, 0.42 – 1.10, p = 0.12). According to these data, the Bayesian posterior predicting model suggested a probability to develop post-spinal arterial hypotension after IVCUS analysis equal to 34.8%; the same probability increased up to 44.6% when patients were analyzed and managed according to PLRT results.

### Secondary endpoints

Regarding fluid administration, all patients were treated by Ringerfundin (B. Braun); the mean total fluid amount was 794 ± 592 ml (Table 3), 925 ± 631 ml for the IVCUS group and 678 ± 529 ml for the PLRT group (p < 0.001, Figure 2). The mean fluid amount given during *pre-anaesthesia phase* was 335 ± 314 ml in the IVCUS group, compared to 169 ± 237 ml in the PLRT group (p < 0.001, Figure 3), while the post-anaesthesia fluid amount resulted identical in both groups (589 ± 370 ml in IVCUS group vs 510 ± 368 in PLRT group, p = 0.075).

**FIGURE 2:**
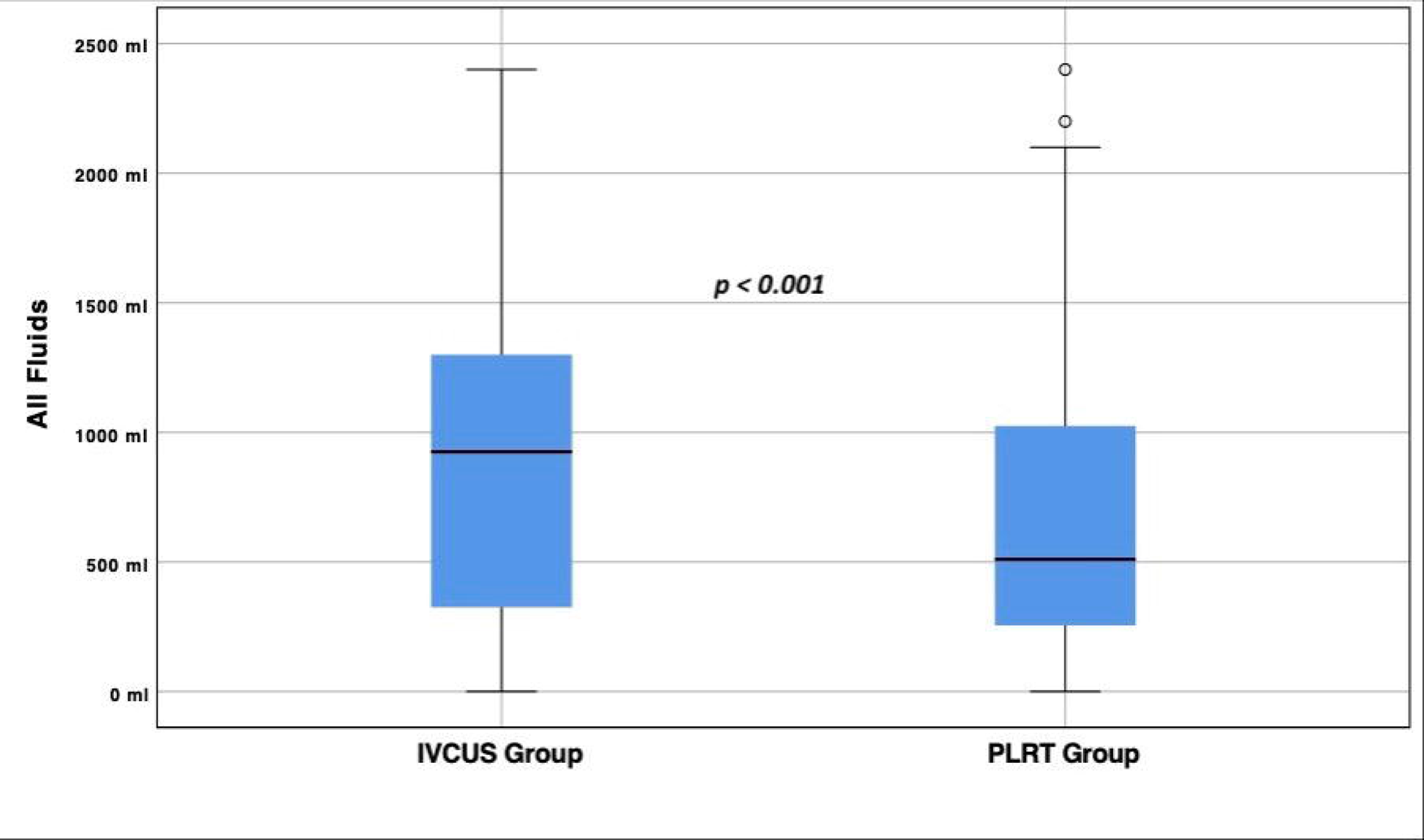
Total fluid amount. Total fluid amount administered to all patients, stratified according to IVCUS and PLRT method; 925 ± 631 ml for IVCUS group and 678 ± 529 ml for PLRT group (p < 0.001).

**FIGURE 3:**
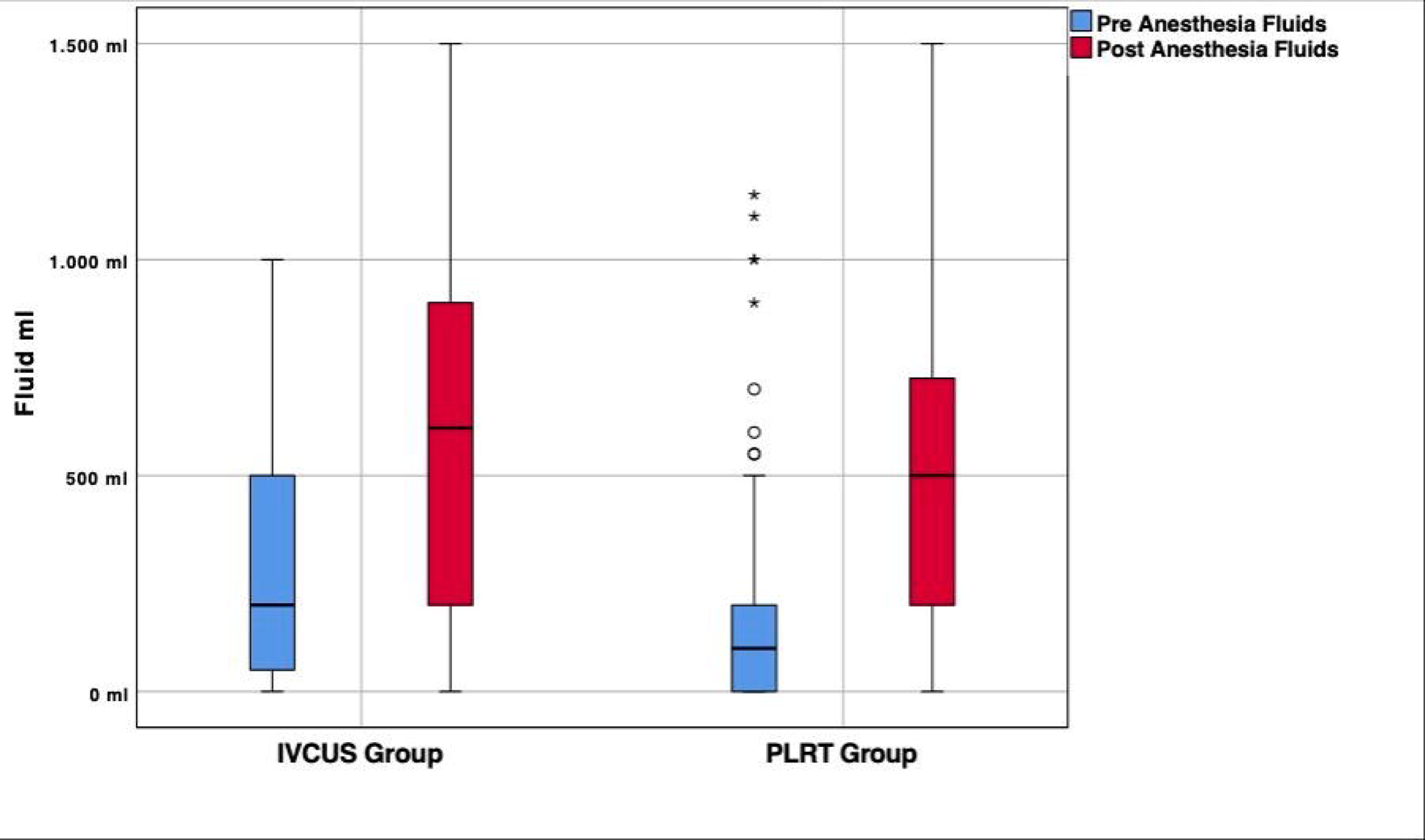
Pre/post-fluid amount distribution. Fluid administration in IVCUS and PLRT group according to pre/post-anaesthesia phase; 335 ± 314 ml in the IVCUS group compared to 169 ± 237 ml in the PLRT group (p < 0.001) in the pre-anaesthesia phase; 589 ± 370 ml in IVCUS group and 510 ± 368 in PLRT group (p = 0.075) during the post-anaesthesia phase.

**TABLE 3:**
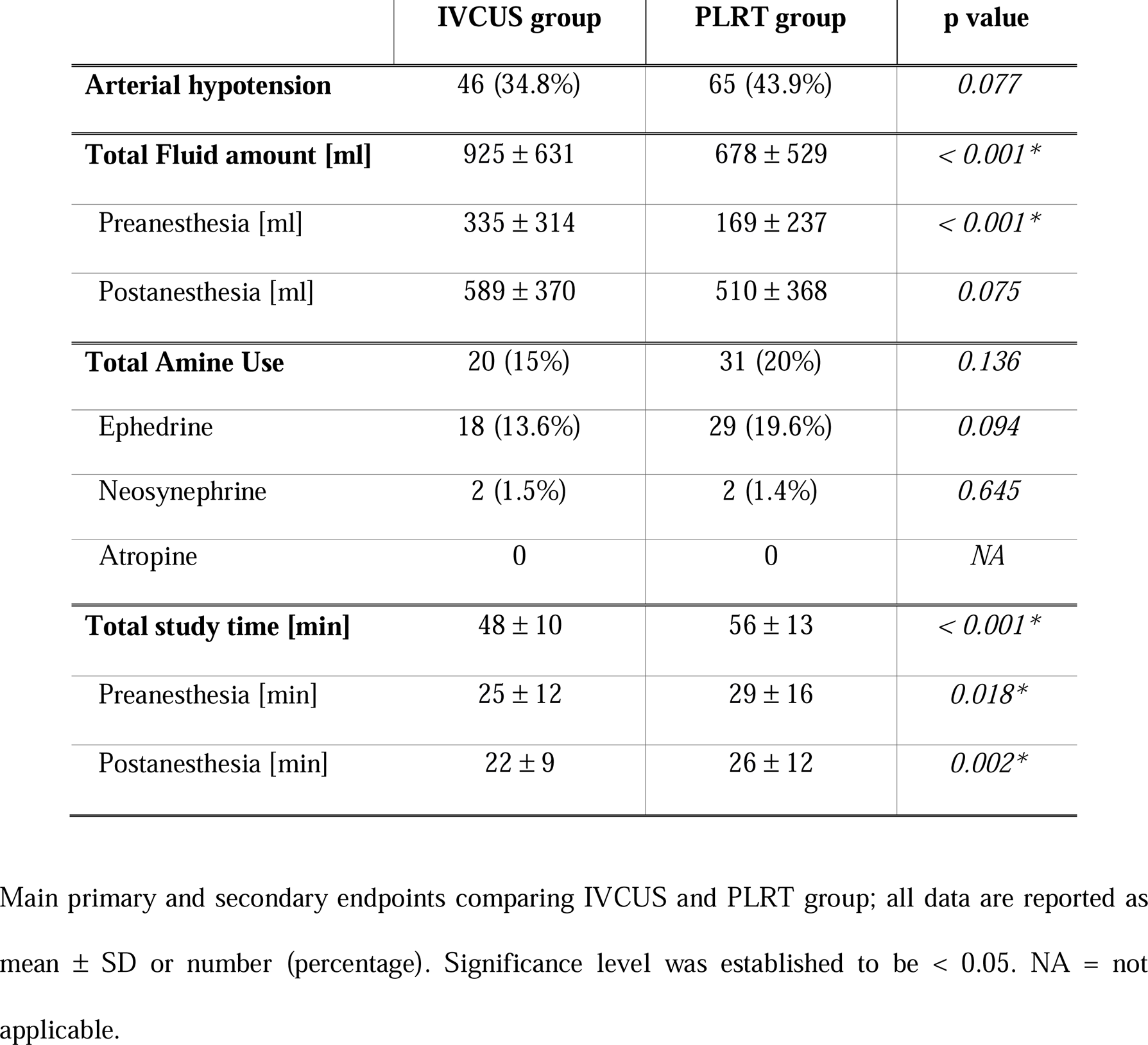
Primary and secondary endpoints

A total of 51 patients (18.2%) required vasoactive support to restore normal arterial pressure, 20 patients (15%) in the IVCUS group and 31 (20%) in the PLRT group (p = 0.136); ephedrine was the drug more used (13.6% in the IVCUS group and 19.6% in the PLRT group, p = 0.094, Table 3). The mean time required to complete the entire procedure was 52 ± 18 min in all patients, 48 ± 10 min in the IVCUS group and 56 ± 13 min in the PLRT group (p < 0.001, Figure 4). Finally, deviation from protocol were detected in 13 (4,6%) cases and led to a drop-out; 2 patients (0.7 %) were given prilocaine for spinal anaesthesia, while 3 patients (1%) developed selective block.

**FIGURE 4:**
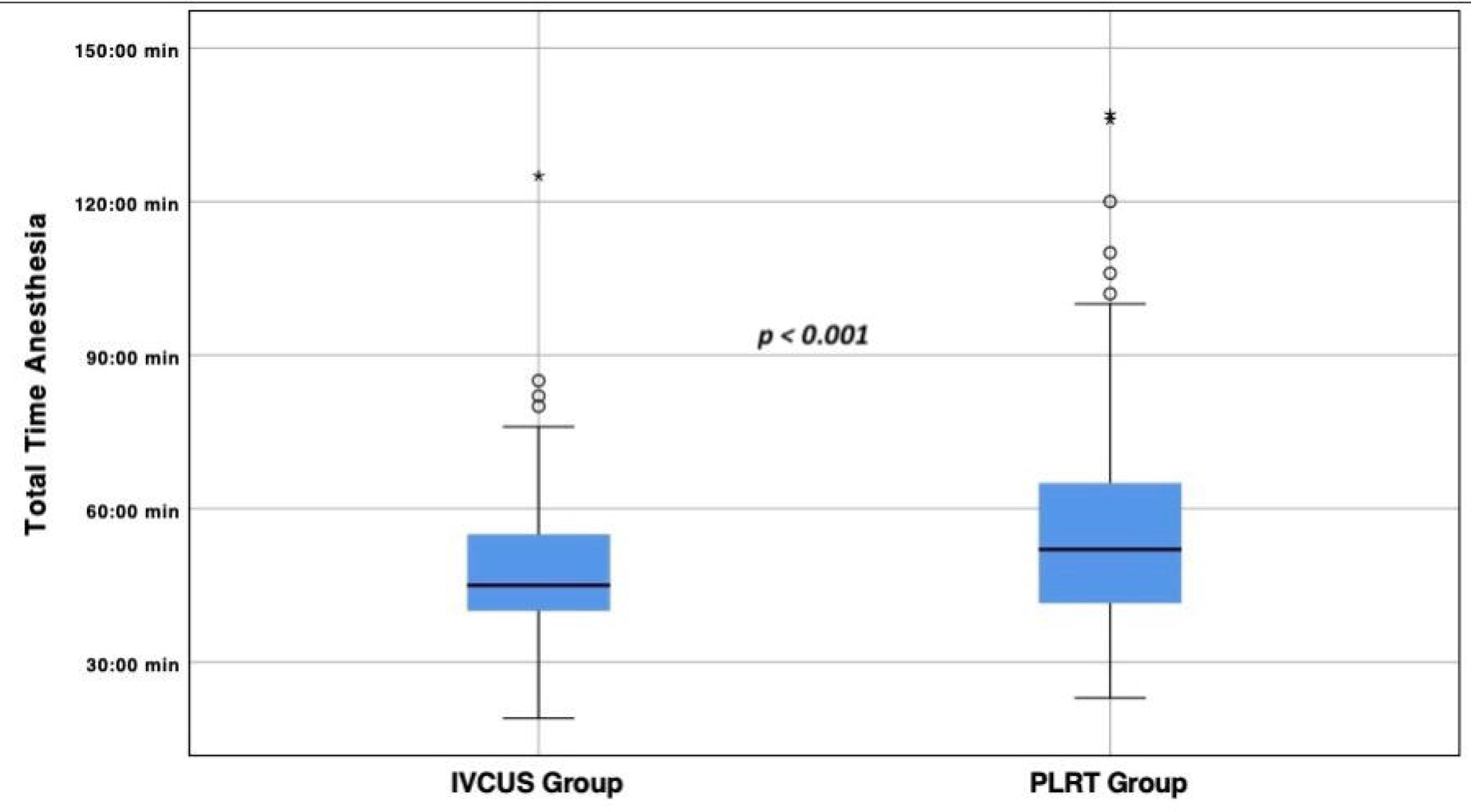
Total time anaesthesia. Total time anaesthesia stratified according to method used for fluid responsiveness assessment; the PLRT group (56 ± 13 min) resulted more time-consuming compared to IVCUS group (48 ± 10 min; p < 0.001).

## DISCUSSION

In critical setting, non-invasive methods to evaluate patients’ volemic status have been of interest during recent years [17]. Fluid responsiveness assessment in spontaneous breathing patients prior to spinal anaesthesia could potentially prevent spinal anaesthesia-induced hypotension, thus ameliorating patients’ outcome [18,19,24,26–32]. PLRT has hitherto been the method of choice in this setting, although literature in favor of IVCUS appears to become more common [23,33,34]. No definite superiority of one method over the other has however been assessed so far. In our post-hoc analysis of the ProCRHYSA trial, the rate of post-spinal arterial hypotension was lower in the IVCUS group in comparison to the PLRT group, even though it resulted in a trend that did not reach statistical significance. This can be in part due to the design of the post-hoc analysis itself, for which a specific power analysis was not performed. Nonetheless, the suggested reduction in the relative risk of post-spinal hypotension through IVC ultrasound instead of PLRT, a test often considered as “standard procedure” in spontaneous-breathing patients, could still influence the every-day practice.

Moreover, the analysis of fluid management also resulted of relevant interest. The IVCUS group received an increased fluid amount before anaesthesia compared to the PLRT group; this fluid administration was not indiscriminate to all IVCUS group patients, but tailored instead to each patient’s haemodynamic status as resulted from the IVCUS assessment. This tailored fluid administration allowed to achieve less arterial hypotension, avoiding at the same time possibly dangerous fluid overload. Although the ProCRHYSA trial was reserved for ASA I-III patients, a similar protocol could in the future be applied to more fragile and unstable patients, given the capability to avoid dangerous fluid overload deriving from this method. This data, associated with a trend towards greater use of amines in the PLRT group, could further suggest a greater efficacy of IVCUS compared to PLRT, even if future validation studies concerning the implementation of portable ultrasound in pre-spinal anaesthesia setting will clarify the role of POCUS in this specific regard.

To date, POCUS applications is rapidly expanding to many clinical fields, included peri-operative medicine; according to recent *World Health Organization* recommendations, it represents one of the 5 priorities to be developed in the field of Critical Care during the next years [43]. The measurement of IVC diameter through the subcostal window is an easy to learn, safe and non-invasive method [44,45]. If routinely applied to fragile patients with cardiovascular comorbidities before anaesthesia, it could not only detect hypovolemic patients benefitting from a pre-procedural volemic repletion, but also guide the titration of fluid therapy, avoiding empiric fluid challenges and the subsequent risk of volume overload and potential heart failure in subjects with a reduced cardiovascular compliance. IVCUS should thus be increasingly considered by anaesthesiologists as a POCUS tool for pre-operative assessment and optimization.

The PLRT method resulted more time-consuming; this aspect may have significant implication, if this test has to be considered for a routine application to high-turnover surgical unit, where anaesthesia-controlled time represents an important part of the total turnover time [46,47]. Although Teboul JL et colleagues identified PLRT as the method of choice in order to predict fluid responsiveness in spontaneous breathing patients [35], others studies draw different conclusions and circumscribe the possible applications of this test. Monnet X. and colleagues [48] state that the high diagnostic sensitivity of PLRT remains confined to the critical care setting, where it should be used in the context of a more global cardiovascular assessment and where a direct measurement of cardiac output can also be performed. In the ProCRHYSA trial, the use of PLRT was not associated to a reduction of the incidence of post-spinal anaesthesia hypotensive episodes compared to the standard method [39].

This analysis presents some limitations. First, it was a post-hoc analysis for which a pre-trial power analysis has not been performed; the limitation of the results’ significance can be mainly due to the study design, and necessarily requires the execution of future studies with greater statistical power to confirm the statistical trend suggested. Moreover, additional studies are also needed to assess whether IVCUS can be implemented to prevent excessive fluid overload in more fragile patients undergoing anaesthesia (i.e. ASA IV-V patients). The addressing of this specific patients’ group demands however a new specifically designed, prospective, randomized, controlled trial. Additional limitations are the same discussed in the ProCRHYSA study [49], especially the impossibility to blind patients to the group allocation, the exclusive involvement of ASA I-III patients and the ultrasound operator-dependency intrinsic to ultrasound evaluations.

## CONCLUSION

Fluid responsiveness assessment in spontaneous breathing patients prior to spinal anaesthesia could potentially prevent post-spinal hypotension, thus ameliorating patients’ outcome; PLRT has hitherto been the method of choice in this setting. In our post-hoc analysis we reported that IVCUS, as a method to guide fluid administration in patients undergoing spinal anaesthesia prior to elective surgery, enables to reduce the incidence of post-spinal hypotension when compared to PLRT. Further studies with greater statistical power are required to determine if the trend shown in our study is confirmed. In this scenario, identifying further ways to implement POCUS in the daily practice could allow ever better patient-tailored approach and management.

## Supporting information

Supplemental Material Consort Check List

## Data Availability

The datasets used and/or analyzed during the current study are available from the corresponding author on reasonable request.

## LIST OF ABBREVIATIONS

ACE-Inhibitors: Angiotensin-converting enzyme
ASA: American Society of Anesthesiologists
CI: Confidence Interval
ESICM: European Society Intensive Care Medicine
etCO2: End-tidal carbon dioxide
IMAO: Monoamine oxidase inhibitors (MAOIs)
IQR: Interquartile Range
IVC: Inferior Vena Cava
IVC-CI: IVC Collapsability Index
IVCUS: Inferior Vena Cava Ultrasound
MAP: Mean Arterial Pressure
PLRT: Passive Leg Raise Test
POCUS: point-of-care ultrasound
ProCRHYSA: PROtocolized Care to Reduce HYpotension after Spinal Anaesthesia
SA: Spinal Anestesia
SSRI: Selective serotonin reuptake inhibitors
SVR: systemic vascular resistance

## DECLARATION

### Ethics approval and consent to participate

The study has been approved by the Ethics Committees (Ref. Cantonal Ethics Committee n. CE2796, international registration number NCT02070276); written informed consent was obtained from patients prior to randomization.

### Consent for publication

Not applicable

### Authors’ contributions

SC conceived the study, designed and coordinated the protocol and recruited patients, furthermore he drafted the manuscript and performed the statistical analysis, AG drafted the manuscript and performed the statistical analysis, MF participated in the design of the study, performed the statistical analysis and drafted the manuscript, ET participated in the draft of the manuscript, GB participated in the statistical analysis and drafted the manuscript, AP participated in the design of the study, performed the statistical analysis and drafted the manuscript, AB performed the statistical analysis and drafted the manuscript, JA participated in the design of the study, performed the statistical analysis and drafted the manuscript, AS participated in the design of the study, coordinated the protocol recruited patients, furthermore he drafted the manuscript and performed the statistical analysis. All authors read and approved the final manuscript.

